# Assessing CT-based Volumetric Analysis via Transfer Learning with MRI and Manual Labels for Idiopathic Normal Pressure Hydrocephalus

**DOI:** 10.1101/2024.06.23.24309144

**Authors:** Meera Srikrishna, Woosung Seo, Anna Zettergren, Silke Kern, Daniel Cantré, Florian Gessler, Houman Sotoudeh, Jakob Seidlitz, Joshua D. Bernstock, Lars-Olof Wahlund, Eric Westman, Ingmar Skoog, Johan Virhammar, David Fällmar, Michael Schöll

## Abstract

**Background:** Brain computed tomography (CT) is an accessible and commonly utilized technique for assessing brain structure. In cases of idiopathic normal pressure hydrocephalus (iNPH), the presence of ventriculomegaly is often neuroradiologically evaluated by visual rating and manually measuring each image. Previously, we have developed and tested a deep-learning-model that utilizes transfer learning from magnetic resonance imaging (MRI) for CT-based intracranial tissue segmentation. Accordingly, herein we aimed to enhance the segmentation of ventricular cerebrospinal fluid (VCSF) in brain CT scans and assess the performance of automated brain CT volumetrics in iNPH patient diagnostics.

**Methods:** The development of the model used a two-stage approach. Initially, a 2D U-Net model was trained to predict VCSF segmentations from CT scans, using paired MR-VCSF labels from healthy controls. This model was subsequently refined by incorporating manually segmented lateral CT-VCSF labels from iNPH patients, building on the features learned from the initial U-Net model. The training dataset included 734 CT datasets from healthy controls paired with T1-weighted MRI scans from the Gothenburg H70 Birth Cohort Studies and 62 CT scans from iNPH patients at Uppsala University Hospital. To validate the model’s performance across diverse patient populations, external clinical images including scans of 11 iNPH patients from the Universitätsmedizin Rostock, Germany, and 30 iNPH patients from the University of Alabama at Birmingham, United States were used. Further, we obtained three CT-based volumetric measures (CTVMs) related to iNPH.

**Results:** Our analyses demonstrated strong volumetric correlations (ρ=0.91, p<0.001) between automatically and manually derived CT-VCSF measurements in iNPH patients. The CTVMs exhibited high accuracy in differentiating iNPH patients from controls in external clinical datasets with an AUC of 0.97 and in the Uppsala University Hospital datasets with an AUC of 0.99.

**Discussion:** CTVMs derived through deep learning, show potential for assessing and quantifying morphological features in hydrocephalus. Critically, these measures performed comparably to gold-standard neuroradiology assessments in distinguishing iNPH from healthy controls, even in the presence of intraventricular shunt catheters. Accordingly, such an approach may serve to improve the radiological evaluation of iNPH diagnosis/monitoring (i.e., treatment responses). Since CT is much more widely available than MRI, our results have considerable clinical impact.

## 1. Introduction

Idiopathic normal pressure hydrocephalus (iNPH) is a treatable neurological condition principally characterized by an enlargement of cerebral ventricles. This condition is associated with a triad of clinical symptoms including gait instability, cognitive impairment, and urinary incontinence^1,2^. A recent scientific discourse suggests the term Hakim disease, but consensus use of this nomenclature has not yet been reached^3^. The standard treatment for iNPH involves neurosurgical diversion of cerebrospinal fluid (CSF) via the placement of an intraventricular shunt^2,4^. The estimated prevalence of iNPH is approximately 1.5 to 3.7% among individuals aged 65 and older, rising to 4 to 6% in those aged 80 and above^5,6^. The work-up of suspected iNPH requires an assessment of structural brain imaging and clinical history consistent with these imaging results^2^. For those who meet these criteria, the definitive diagnosis often involves a lumbar puncture/CSF drainage, complemented by serial clinical assessments at a specialized center^7,8^. While less invasive than intracranial shunting, lumbar punctures can still entail risk, albeit rare, for headaches and discomfort^9^.

Typical morphological features of iNPH are most often assessed using magnetic resonance imaging (MRI), focusing on radiological markers such as the Evans’ index^10^, callosal angle^11,12^, and disproportionately enlarged subarachnoid spaces^13^. The Evans’ index is defined as the ratio of the maximal width of the frontal horns of the cerebral ventricles to the maximum inner diameter of the skull. The callosal angle, which measures the angle between the lateral ventricles in a coronal image perpendicular to the bi-commissural plane, is another critical radiological marker described in clinical guidelines for diagnosing iNPH and assessing ventricular enlargement in the context of potential cerebral atrophy^2^. Importantly, these measurements rely on assessments made in the bi-commissural plane^14^ and while the Evans’ index provides insight into the extent of hydrocephalus by measuring the degree of ventricular expansion, it lacks specificity among the elderly population as it does not provide information about cerebral atrophy^15,16^. Truly volumetric measurements offer a more direct method for assessing ventricular size, overcoming previous technical limitations in clinical practice. In line with this, research has shown that three-dimensional analysis of ventricular volume, achieved through semi-automated volumetric techniques or atlas-based segmentation, is more sensitive than the Evans’ index for monitoring ventricular size changes after shunt treatment in patients with iNPH^17–19^.

Ventricular volume measurement is typically performed using automated or manual segmentation of ventricular cerebrospinal fluid (VCSF) in research environments. MRI, known for its superior soft-tissue contrast, facilitates the precise identification of the bi-commissural plane and other crucial anatomical landmarks^20^, making it the preferred method for assessing diagnostic metrics related to iNPH and for the automated volumetric quantification of ventricles and other brain tissue types^21–24^. Conversely, computed tomography (CT) offers a more accessible and cost-effective alternative, with the advantages of shorter scan durations, broader availability, and the absence of strong magnetic fields, making it particularly valuable for patients who are unable to undergo MRI examinations. Moreover, CT scans are routinely used in the clinical assessments of traumatic brain injury and other neurodegenerative disorders. The frequent use of CT offers the opportunity to detect incidental morphological changes related to iNPH, potentially enabling earlier diagnosis and subsequent treatment of the condition^25^. Despite these advantages, current radiological assessments of CT still largely depend on subjective visual rating methods that are time-consuming, labor-intensive, require significant expertise, and are highly susceptible to variability in inter-rater reliability^1,2^.

Interestingly, while numerous studies have successfully demonstrated brain tissue segmentation and volumetric assessment using MR images^26,27^, these techniques have encountered challenges when attempted using CT scans with inferior soft tissue contrast. Recent research has thus explored robust automated and semi-automated segmentation(s)^28,29^ and volumetric assessments of CT images using state-of-the-art image processing techniques centred on deep learning; specifically, segmentation models employing deep learning architectures, such as fully connected convolutional neural networks and 2D/3D U-Nets have been utilized^30–33^. However, most deep-learning-based studies have thus far been conducted using limited datasets and relied on manually annotated structural labels. In our previous research, we successfully employed MR-based tissue class segmentations, derived automatically using established and validated methods, to train deep-learning-models for tissue classification in brain CT scans^34^. To accomplish this, we trained U-Net-based deep-learning-models to segment grey matter, white matter, CSF, and intracranial volume (ICV) in brain CT images with slice thicknesses ranging from 3-5 *mm*^34^. Leveraging our expertise and recognizing the underused potential of CT in diagnosing and managing neurological disorders such as iNPH globally, our current goal was to develop deep-learning-models trained on both manually and automatically derived labels. These models were designed to segment VCSF, specifically targeting the lateral ventricles in brain CT images, using transfer learning techniques that are optimal for scenarios with limited training data^35,36^.

Furthermore, we aimed to extract CT-based volumetric metrics (CTVMs) related to iNPH through deep learning, and to evaluate their performance in differentiating iNPH patients from controls, as well as to quantify VCSF volume before and after shunt surgery in iNPH patients as a means to automatically monitor treatment efficacy. We ultimately determined three volumetric ratios: the lateral ventricular volume to intracranial volume (VCSF/ICV), also known as the 3D Evans’ index (3D-EI), which offers a three-dimensional enhancement of the traditional Evans’ index with potentially increased sensitivity and precision; the brain volume to intracranial volume, occasionally referred to as the brain parenchymal fraction (BV/ICV), which indicates total parenchymal atrophy; and the ventricular volume to CSF volume (VCSF/CSF), providing a more direct measure of hydrocephalus. We propose that these volumetric parameters may therefore enhance our ability to use CT images clinically for the rapid and objective detection of hydrocephalus and ultimately other neurodegenerative disorders.

## 2. Materials and Methods

We developed models based on deep learning capable of segmenting lateral VCSF from brain CT images. These models were trained using a transfer learning approach, incorporating both automatically derived MR labels and manually derived CT labels. The model was developed in three stages: pre-processing, training the deep-learning-model with MR-based labels, and training the deep-learning-model with manually derived CT labels. In this pipeline, we initialized the latter model using transferable features from the MR-VCSF label-trained pre-trained model. **Figure 1** provides an overview of the model development pipeline.

**Figure 1.**
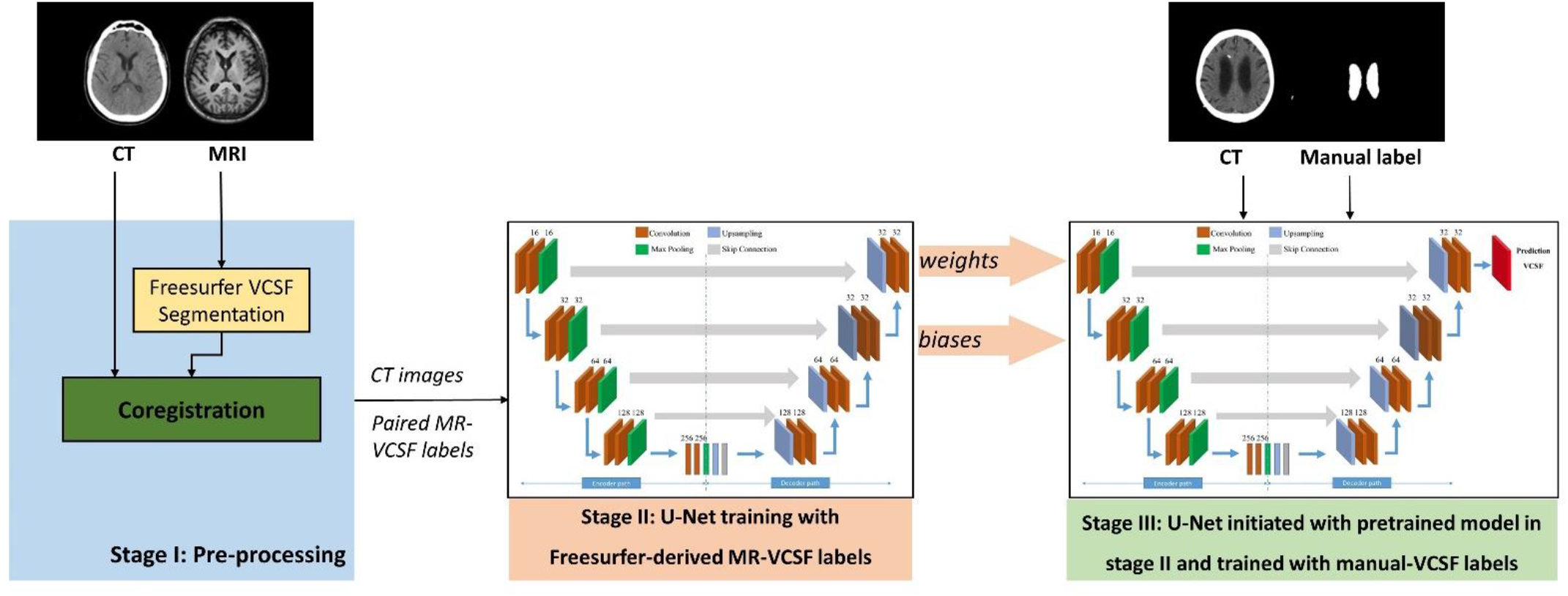
Overview of model development and training. Stage I: In the pre-processing stage, 734 CT-MRI pairs of 70-year-old individuals from the Gothenburg H70 Birth Cohort Studies were coregistered. MR images were segmented to lateral VCSF using FreeSurfer. Stage II: Paired CT- and MR-VCSF labels were split into training, validation, and unseen test datasets for a three-fold cross-validation. A 2D U-Net-based deep-learning-model was developed and trained to predict lateral VCSF with paired CT images and MR labels as training inputs. Stage III: The weights and biases from Stage II deep-learning-models were used to initialize another U-Net-model, which was further trained with paired CT and manual CT-VCSF labels from 62 iNPH patient datasets.

### Datasets

#### Gothenburg H70 Birth Cohort Studies

The datasets used for training and validating the models obtained through deep learning (via MR-derived labels) were obtained from the predominantly healthy participants within the Gothenburg H70 Birth Cohort Studies^37^. These multidisciplinary longitudinal epidemiological studies involve six birth cohorts, each initially enrolled and examined at the age of 70, to investigate the ageing population in Gothenburg, Sweden. Data was collected from 2014 to 2016. The H70 study was approved by the Regional Ethical Review Board and by the Radiation Protection Committee in Gothenburg, Sweden. Brain imaging was conducted at Aleris Röntgen Annedal in Gothenburg (Aleris Healthcare AB, Stockholm, Sweden). CT images were available for *n* = 917 participants of which the vast majority (99%; *n* = 904) were cognitively normal. Of these 917 participants, 79% (*n* = 744) underwent MR scanning within a day from their respective CT scan; for model development, we included these ∼same-day acquisitions of CT and MR images from the respective 744 participants (52.6*%* female, 70.44 ± 2.6 *years*, cognitively normal, *n* = 722). CT scanning was conducted using a 64-slice Philips Ingenuity CT system (Philips Medical Systems, Best, Netherlands); resultant images had a slice thickness of 0.9 *mm*, an acquisition matrix of 512 × 512 and voxel size 0.5 × 0.5 × 5.0 *mm*^*3*^. MR scanning was conducted on a 3-Tesla Philips Achieva system (Philips Medical Systems). The protocol included the acquisition of T1-weighted images with a field of view of 256 × 256 × 160, voxel size: 1 × 1 × 1 *mm*^*3*^, echo time: 3.2 *ms*, repetition time: 7.2 *ms*, flip angle: 9*°*.

#### Uppsala University Hospital

For the deep-learning-models trained with manually annotated labels, this dataset included 62 CT scans from 33 iNPH patients (79.3 ± 6.46 *years*, 51*%* female) at Uppsala University Hospital. Manual labels of the lateral ventricles were curated for the CT scans. Out of all patients, 23 had post-shunt scans available that featured intraventricular catheters. Manual segmentation was performed by three raters under the guidance of a board-certified neuroradiology specialist (DF).

#### Clinical Validation Datasets

To validate the diagnostic performance of the CT-based volumetric measures developed using deep learning, an external dataset was assembled. It included 11 iNPH CT scans from Universitätsmedizin Rostock (74.5 ± 7.39 *years*, 21*%* female) and 30 iNPH scans from the University of Alabama at Birmingham (70.4 ± 9.1 *years*, 33.33*%* female).

### Model Development

#### Pre-processing

Images were pre-processed using SPM12 (http://www.fil.ion.ucl.ac.uk/spm, MATLAB 2023a). All CT, MR images, and CT-derived manual segmentations were converted to NIfTI format(s). In the Gothenburg H70 Birth Cohort, the images were visually assessed, and the AC-PC planes were aligned. VCSF segmentations were derived from MR images using Freesurfer 5.3.0^38^ processed through theHiveDB system^39^. To train the model in the CT space, the MR labels were co-registered to CT images in SPM12^40^ using rigid body transformation. The registration between output CT and MR labels of each dataset was visually assessed; ten datasets were found to have faulty co-registrations and were discarded.

### Model Development Using Automatically Derived MR Labels

At this stage, the deep-learning-models were trained using MR-VCSF labels. The 734 pre-processed datasets from the Gothenburg H70 Birth Cohort consisting of CT images, and Freesurfer-derived MR-VCSF labels were subdivided into training and cross-validation groups. We used a three-fold cross-validation method for our analysis. From a pool of 734 datasets, we randomly divided them into three equal parts. For each cycle of validation, one part was designated as the unseen test set, and the remaining two were merged and further split into 400 training and 100 validation sets. We trained the model parameters using the training sets and fine-tuned them with the validation sets. The effectiveness of the model was then evaluated using the test sets, which had not been seen by the model during training. The U-Net-based deep-learning-model used at this stage was developed in Python 3.10.14, using TensorFlow 2.10.0 and Keras 2.10.0. The models were trained on an MSI GeForce RTX 2080 Ti, 11GB RAM graphical processing unit (GPU). The CT images were provided as training inputs, and the MR labels were assigned as training labels. The U-Net was designed to accept 2D slices of size 512 × 512, from the axial plane of the input brain scan. Once the datasets were organized, we constructed a model with 1,177,649 trainable hyperparameters. We used a batch size of 16, incorporating features such as early stopping and automatic reduction of the learning rate based on the training progress. The model underwent training for 50 epochs. For semantic segmentation, we utilized binary cross entropy and the Dice coefficient as loss functions to optimize the model’s performance. We employed the adaptive moment estimator (Adam) optimizer with a learning rate of 0.00001 to estimate these parameters. All weights were initialized using a normal distribution with a mean of 0 and a standard deviation of 0.01, while all biases were set to zero.

### Model Development Using Manually Derived CT-VCSF Labels

In the final stage of the model development, the U-Net-based deep-learning-models were trained with manual labels derived from CT images. The 62 datasets from the Uppsala University Hospital consisting of paired CT images, and manual lateral CT-VCSF labels were subdivided into training and cross-validation groups for a three-fold cross-validation. During each training fold, the 62 paired CT and manual CT-VCSF labels were randomly categorized into training (*n* = 32), testing (*n* = 20), and validation (*n* = 10), groups. To reduce overfitting, we increased the training and validation group sizes using data augmentation^41^. The U-Net models were developed in Python 3.10.14, using TensorFlow 2.10.0 and Keras 2.10.0 and trained on an MSI GeForce RTX 2080 Ti, 11GB RAM graphical processing unit. We incorporated the U-Net, previously trained with automatically derived MR-VCSF labels, by initializing the encoders of the current U-Net models. All other parameters and training conditions remained consistent with our previous models. After adjusting the trainable parameters, the model underwent training propagations within each epoch. These propagation algorithms iterated for all epochs, with parameters updated accordingly. Training ceased upon model saturation or completion of the specified number of epochs. The resulting models were then preserved for the automated VCSF segmentation of CT scans, leveraging features acquired from both automatically derived MR labels and manually derived CT labels.

### Image Processing and Analysis

#### CT-based Volumetric Metrics (CTVMs)

Deep-learning-based CT-VCSF segmentation was executed in Python 3.10.14, using TensorFlow 2.10.0 and Keras 2.10.0. The model’s predictions were independent of MRI or any other label or user input, and pre-processing of CT images. The CT images from all cohorts served as inputs to the trained U-Net models. As we used cross-validation, we were able to effectively use the same data (from Gothenburg H70 Birth Cohort studies and Uppsala University Hospital datasets) for both training and testing our models. Cross-validation allowed us further to repeatedly train our model on different subsets of the data and then test it on the remaining portions, instead of simply splitting this data into a training set and a separate test set^42^. Hence, we could use the training inputs as test datasets by applying models to data unseen during the training process. The models efficiently generated lateral VCSF segmentation maps from CT images using the trained hyperparameters, completing the task in approximately 10 seconds per dataset. Additionally, we derived grey matter, white matter, ICV, and CSF maps using U-Net-based deep learning models trained in our previous study^34^.

The resulting segmentation maps represent probability or confidence maps, with each pixel indicating the likelihood of belonging to the VCSF labels. These maps underwent binarization using global thresholding set at 0.5. Stacking the slices of segmentation maps produced a 3D image, from which the sum of voxels was calculated and then multiplied by the voxel size to determine brain tissue volume (in mL). Total brain volume (BV) was derived from the binarized parenchymal maps. By using the lateral VCSF, CSF, ICV, and BV segmentation maps obtained from CT images via deep-learning-models, we derived the following iNPH-related CT-based volumetric measures: VCSF/ICV (3D-EI), BV/ICV, and VCSF/CSF.

#### Ground Truth Label Volumes

Manually derived CT-VCSF labels and automatically derived MR-VCSF labels were binarized using global thresholding set at 0.5, and volumes were quantified by the sum of voxels, multiplied by the voxel size.

### Statistical Analysis

Shapiro–Wilk test was used to examine the Gaussian distribution of the continuous volumetric metrics (*p* > 0.05). The volumetric similarity between automatically derived CT-VCSF and manually derived CT-VCSF in the Uppsala University Hospital datasets, and CT-VCSF and MR-VCSF in the Gothenburg H70 Birth Cohort was assessed using Spearman rank correlation tests, and the agreement was visualized using Bland–Altman plots with 95% limit of agreement. We also computed and compared the Spearman rank correlation between manually and automatically derived CT-VCSF volumes in pre- and post-shunt procedure in the Uppsala University Hospital datasets. We compared the distribution of CTVMs between the iNPH patients from Uppsala University Hospital and controls from Gothenburg H70 Birth Cohort using the Mann–Whitney test (*p* < 0.05). The accuracy of CTVMs in distinguishing iNPH patients from cognitively normal individuals was assessed by measuring the area under the receiver operating characteristic curve (ROC-AUC) with 95% confidence intervals (CI). Additionally, we conducted a comparison of the distribution of CTVMs between iNPH patients and controls, as well as ROC-AUC analysis, on iNPH patients from the validation clinical datasets and Gothenburg H70 Birth Cohort. All statistical analyses and visualization curation were performed using R version 4.3.2 (2023).

## 3. Results

**Table 1** lists the iNPH-related CT-based volumetric measures for all participants in all cohorts. As hypothesized, the mean CT-VCSF/ICV (3D-EI) was higher among the iNPH patients (0.11 ± 0.05) from the validation iNPH patient datasets, as compared to the predominantly healthy Gothenburg H70 Birth Cohort participants (0.04 ± 0.01). Also, the CT-BV/ICV was lower in the iNPH patients (0.65 ± 0.11) in comparison to cognitively normal individuals (0.74 ± 0.03) from the Gothenburg H70 Birth Cohort.

**Table 1.**
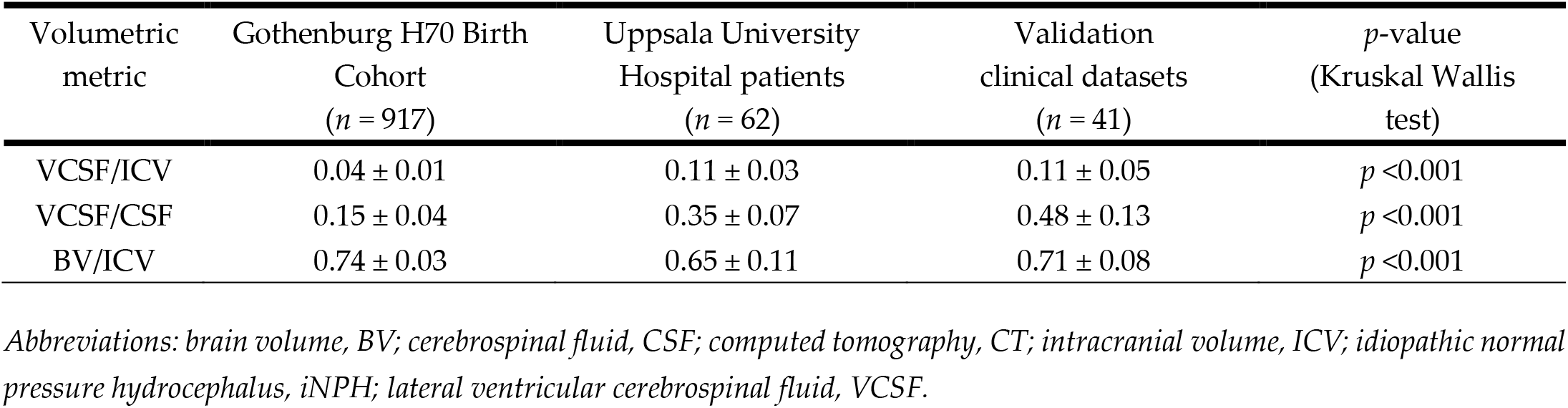
Summary of iNPH-related CT-based volumetric measures obtained through deep learning.

### Segmentation performance compared with manually derived CT-VCSF labels

**Figure 2** illustrates the outputs from our deep-learning-models, showcasing the predicted CT-VCSF maps alongside their corresponding manual CT-VCSF labels for comparison. In the iNPH patient datasets from the Uppsala University Hospital, automatically derived CT-VCSF volumes demonstrated strong correlations with the manually derived CT-VCSF volumes (*ρ* = 0.91, *p* < 0.001; **Figure 3a**, left). The Bland-Altman analysis performed between these two measures yielded a bias of 33.44 mL (24.11*%*) and a standard deviation (SD) of 19.27 *mL* (**Figure 3a**, right) indicating an overestimation through the automatically derived CT-VCSF volumes as compared to manually derived CT-VCSF volumes. Additionally, in the Gothenburg H70 Birth Cohort, CT-VCSF volumes obtained through deep learning demonstrated a strong correlation with the MR-VCSF volumes (*ρ* = 0.91, *p* < 0.001) (**Figure 3b**, left) with Bland-Altman analysis indicating a bias of 4.53 mL (14.9*%*) and SD of 7.31 *mL* (**Figure 3b**, right).

**Figure 2.**
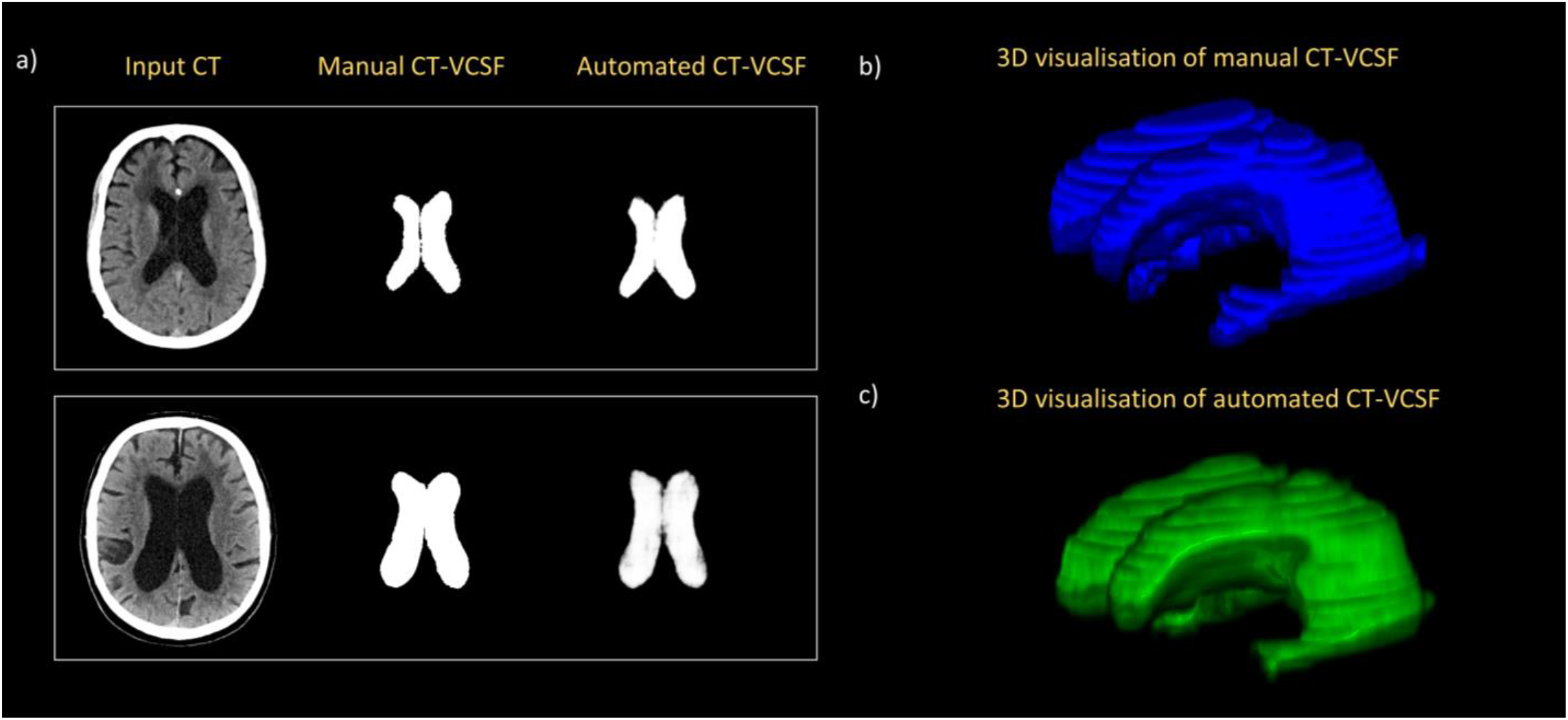
Model outputs. a) Input CT images, manual lateral VCSF, and U-Net predicted lateral VCSF maps of two representative test iNPH patient datasets; b) and c) 3D visualization of manually derived and automatically derived CT-VCSF. *Abbreviations: CT, computed tomography; lateral ventricular cerebrospinal fluid, VCSF*.

**Figure 3.**
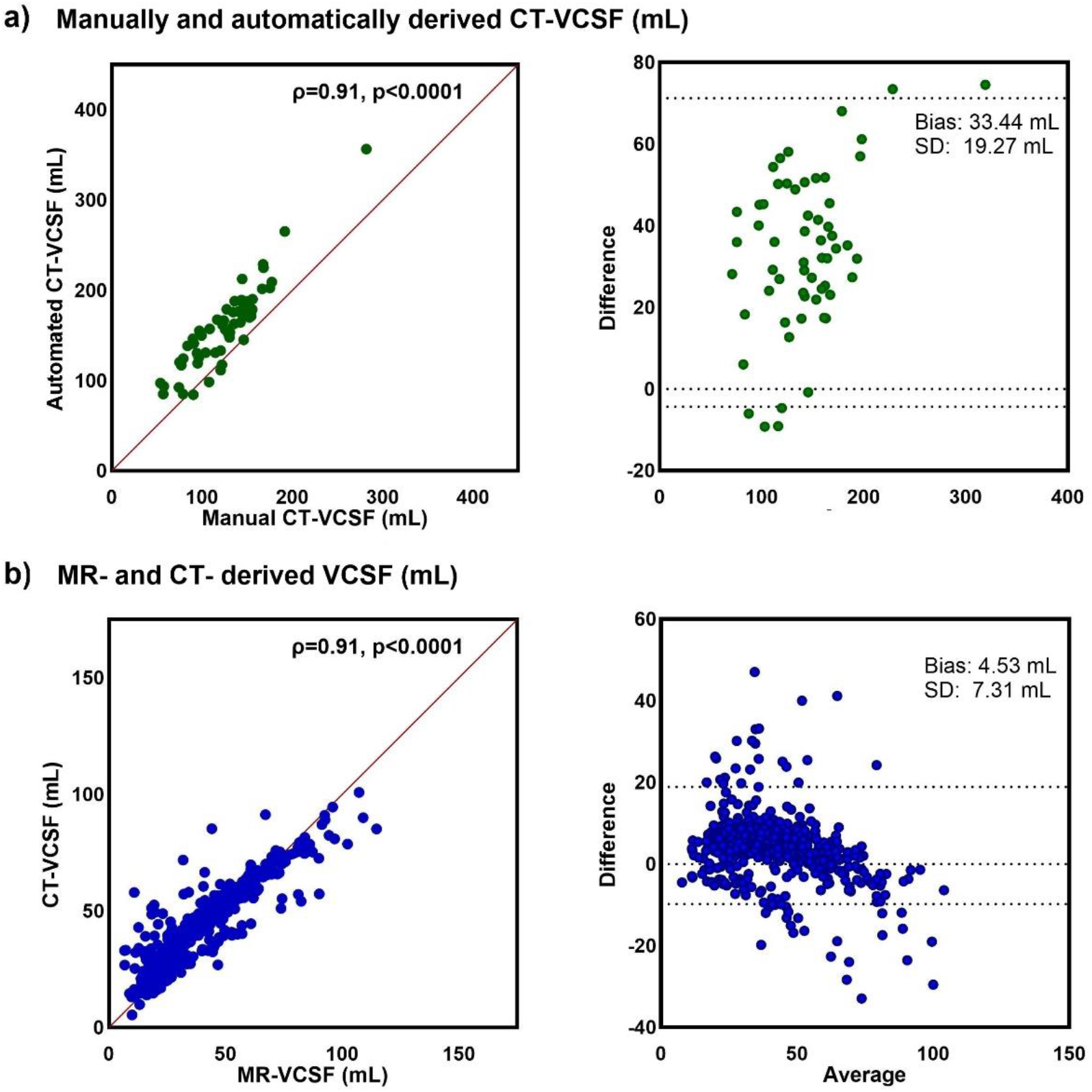
Segmentation performance for volumetry. a) Correlation plots and Bland-Altman analysis plots between manually derived and automatically derived CT-VCSF; b) Correlation plots and Bland-Altman analysis plots between automatically derived MR- and CT-VCSF. *Abbreviations: CT, computed tomography; MRI: magnetic resonance imaging; SD: standard deviation; lateral ventricular cerebrospinal fluid, VCSF*

### Segmentation performance in the presence of shunt

The trained deep-learning-models effectively predicted segmentation maps, even in the presence of intraventricular catheters. **Figure 4a** specifically displays the deep-learning-model’s CT-VCSF predictions at the site of the intraventricular catheter; both pre-and post-shunt VCSF volumetric differences were similar between manual segmentations and those produced automatically by the model, as illustrated in **Figure 4b**.

**Figure 4.**
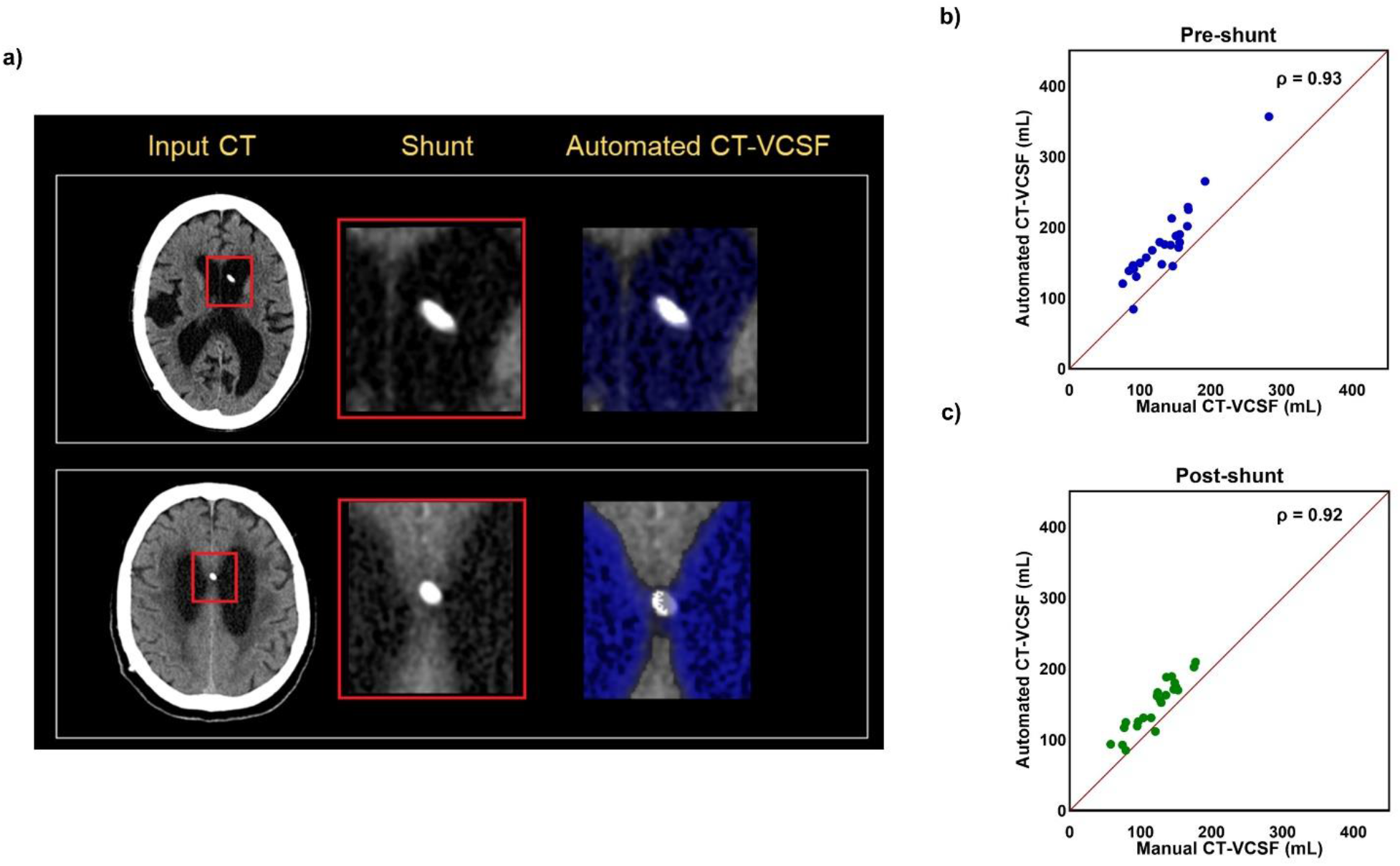
Model predictions with an intraventricular catheter. a) Input CT images with shunts, and U-Net predicted VCSF maps of two representative post-shunt iNPH patient test datasets. On the right, correlation plots between manually derived and automatically derived CT-VCSF volumes of b) pre-shunt and c) post-shunt procedure volumes of iNPH patient datasets (*n* = 23) are depicted. *Abbreviations: CT, computed tomography; lateral ventricular cerebrospinal fluid, VCSF*.

### Diagnostic Performance of CTVMs

In the clinical validation datasets, iNPH-related CTVMs demonstrated high accuracy in distinguishing iNPH patients from controls (datasets from the Gothenburg H70 Birth Cohort), achieving an AUC of 0.97 (95% CI: 0.94 to 1.00) for CT-VCSF/ICV (3D-EI), as shown in **Figure 5a**. However, this was not the case for CT-BV/ICV. Overall, CTVMs effectively differentiated between the two groups, with average CT-VCSF/ICV (3D-EI) (**Figure 6a**) and CT-VCSF/CSF (**Figure 6b**) values being higher in iNPH patients compared to controls.

**Figure 5.**
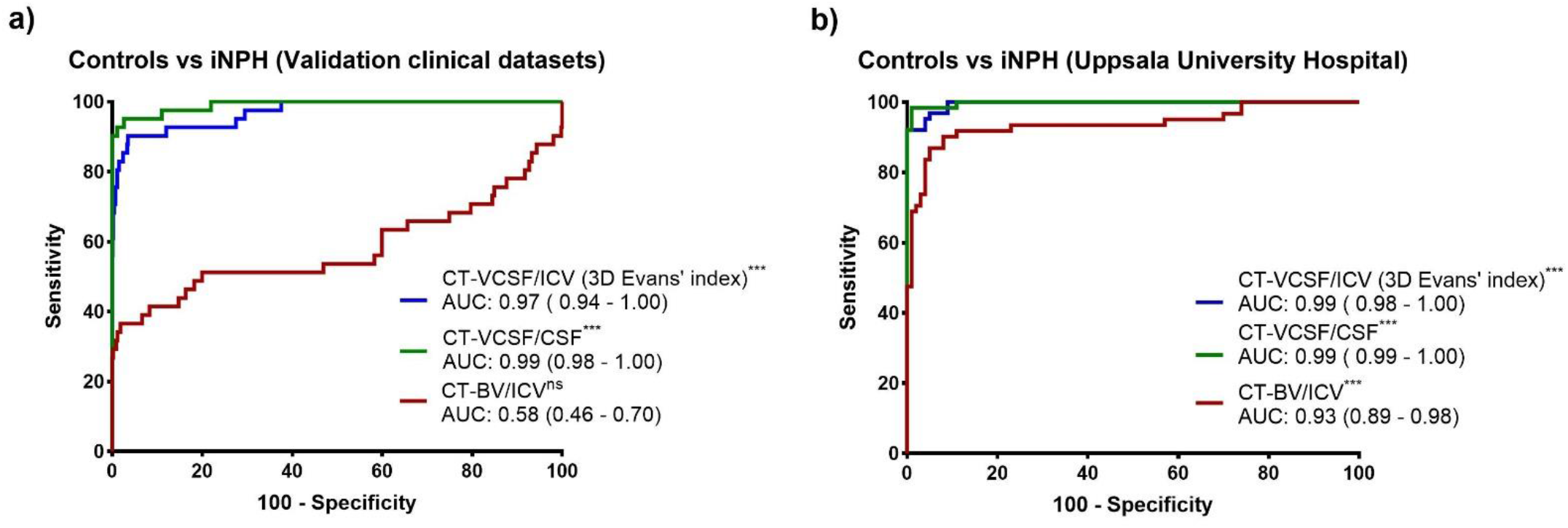
ROC curves for distinguishing iNPH from controls. The plot shows the ROC curves of various CTVMs in distinguishing iNPH patients of a) validation clinical datasets (*n* = 41) and b) Uppsala University Hospital (*n* = 62) from controls obtained from the Gothenburg H70 Birth Cohort (*n* = 917). \*\*\**p* < 0.001; ^ns^not significant. *Abbreviations: brain volume, BV; cerebrospinal fluid, CSF; intracranial volume, ICV; idiopathic normal pressure hydrocephalus, iNPH; lateral ventricular cerebrospinal fluid, VCSF*.

**Figure 6:**
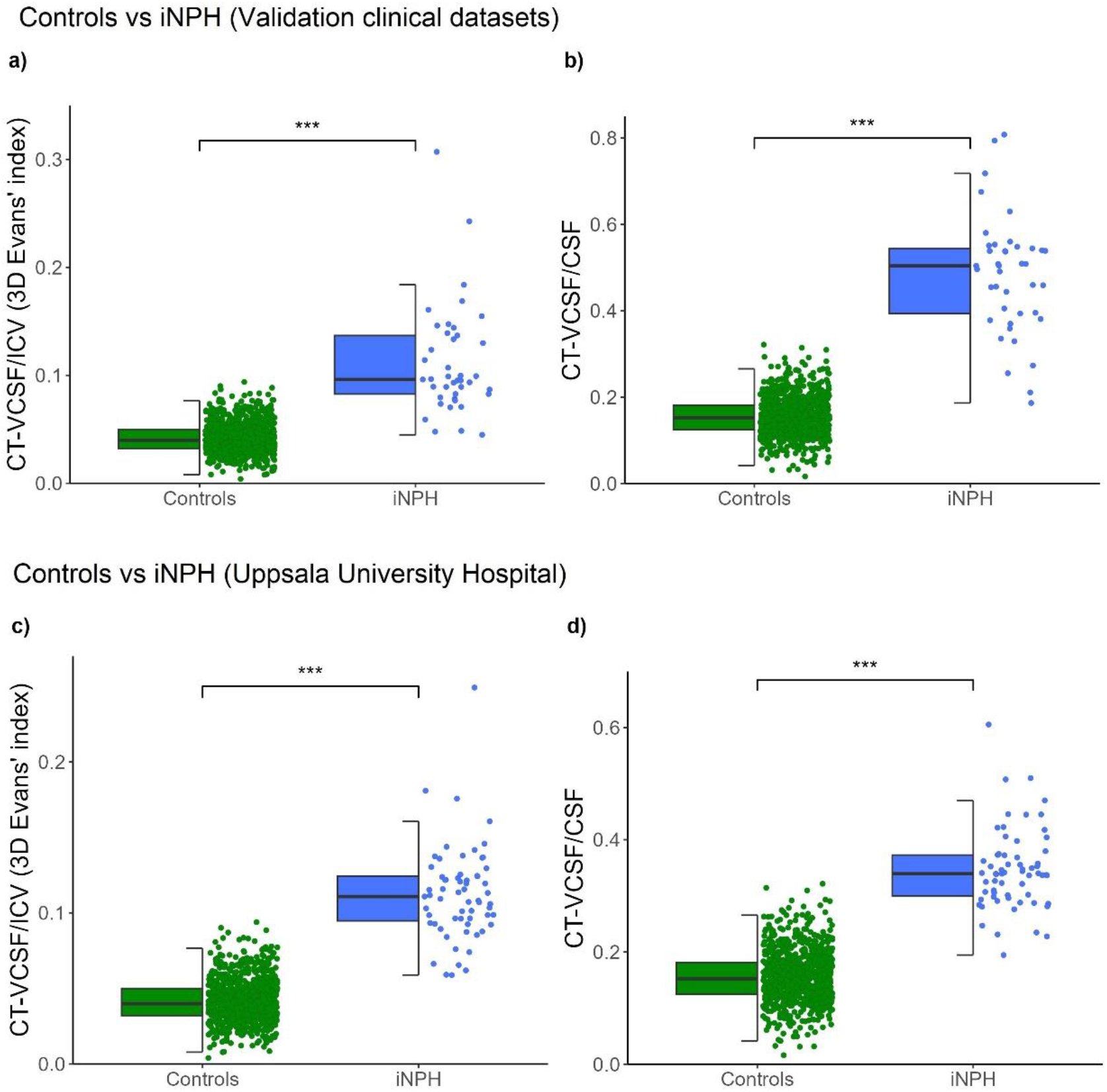
Distribution of CT-based volumetric measures across iNPH patients from a) Validation clinical datasets (*n* = 41) and b) the Uppsala University Hospital (*n* = 62) and controls from Gothenburg H70 Birth Cohort (*n* = 917). \*\*\**p* < 0.001, p values are from the Mann-Whitney test. *Abbreviations: brain volume, BV; cerebrospinal fluid, CSF; computed tomography, CT; intracranial volume, ICV; idiopathic normal pressure hydrocephalus, iNPH; lateral ventricular cerebrospinal fluid, VCSF*.

Additionally, in the Uppsala University Hospital datasets, CTVMs also demonstrated excellent accuracy in distinguishing iNPH patients from controls with an AUC: 0.99, 95% CI: 0.98, 1.00 for CT-VCSF/ICV (3D-EI) (**Figure 5b**). Similar to the validation clinical datasets, CTVMs significantly differentiated between the two groups, with mean CT-VCSF/ICV (3D-EI) (**Figure 6c**), and CT-VCSF/CSF (**Figure 6d**) higher in iNPH patients as compared to controls.

## 4. Discussion

Computed tomography (CT) scans are the major routine method for evaluating brain morphology in both developed and developing countries but typically rely on subjective visual inspection and ratings^43,44^. Adopting reliable automated and quantitative methods, such as machine learning for ventricular segmentation, could substantially improve the diagnostic performance of CT-derived data through enhanced accuracy and consistency, reducing inter-rater variability, and providing substantial benefits in both research and clinical contexts. Most machine learning applications for CT scans still rely on time-consuming, labor-intensive manual labelling, which introduces subjectivity and limits the availability of training datasets. Our study introduces a pioneering approach using deep learning, trained with both automatically and manually derived brain labels; the standard for evaluating iNPH in research and clinical settings. We utilized automated labels from MRI to enhance training efficiency and address the scarcity of annotated data, which poses significant challenges in brain morphology assessment and quantification. By leveraging transfer learning and automated labels, we aim to improve the accuracy and robustness of our models, mitigating the limitations of manual annotation. Once trained, these models operated solely on CT data, eliminating the need for MRI inputs and/or visual assessments. We successfully applied these models to segment ventricular structures from CT images across two distinct diagnostic groups: a predominantly cognitively normal elderly population (*n* = 917) and iNPH patients (*n* = 94), utilizing data from four different sources, thereby demonstrating the method’s robustness and generalizability.

A key finding from our study is that deep learning models, when properly optimized and trained with both automatically and manually derived labels, are highly effective at segmenting VCSF from head CT images with different slice thicknesses (ranging from 3 to 5 *mm*). Automatically derived CT-VCSF labels strongly correlated with manually derived CT-VCSF (*ρ* = 0.91, *p*<0.001). However, the bias of 33.44 *mL* (24.11*%*) in the Bland-Altman analysis suggests a systematic difference between the two measures, with automatically derived CT-VCSF volumes consistently overestimating lateral ventricular volumes in comparison to manually derived CT-VCSF. The systematic overestimation could potentially enhance the distinction between iNPH patients and controls. Indeed, the CT-VCSF-based measures derived automatically demonstrated a high level of accuracy in differentiating iNPH from control subjects. Additionally, the automatically derived CT-VCSF measurements showed a strong correlation with automatically derived MR-VCSF, with a bias of 4.53 *mL* (14.9*%*) and a standard deviation of 7.37 *mL*, indicating reasonable agreement albeit with some variability. Congruent with the literature, our segmentation performance was in line with previous work (i.e., that of Zhou et al. 2021^32^ and Huff et al. 2019^30^). Previously, we confirmed the segmentation similarity between CT brain tissue classes obtained using deep learning and their corresponding MR brain tissue classes determined through established automated segmentation tools^34^. In this study, we further validated the similarity of CT volume segmentations performed by deep learning against manually derived CT volumes, which currently and still serve as the gold standard.

Another critical finding of clinical importance from our study was that CT-based volumetric metrics demonstrated high accuracy in distinguishing iNPH patients from cognitively normal individuals in the H70 Cohort, with AUC values of 0.97 and 0.99 for two volumetric ratios: CT-VCSF/ICV (3D-EI) and CT-VCSF/CSF. This was particularly evident in the validation of clinical datasets. In a review by He et al. 2020^45^, the AUC of iNPH *vs* cognitively normal for manual CT-derived Evans’ index and anteroposterior diameter of the lateral ventricle index (ALVI) were reported to be 0.95 and 0.99, respectively. Compared to MR-derived z-Evans’ index (AUC, 0.91), Evans’ Index (AUC, 0.84) and callosal angle (AUC, 0.93) in Yamada et al. 2015^46^, CT-based volumetric measures exhibited higher diagnostic accuracies. Additionally, in Neikter J et al. 2020^47^, the diagnostic accuracy of MR-derived brain volume ratio (AUC, 0.98), callosal angle (AUC, 0.97), Evans’ Index (AUC, 0.96), and presence of disproportionately enlarged subarachnoid spaces (AUC, 0.96) was comparable to CTVMs. Kockum et al. 2020^48^ developed the INPH Radscale; a radiological assessment tool to quantify ventriculomegaly and other brain changes for diagnosing iNPH. CTVMs also exhibited comparable diagnostic accuracies to iNPH Radscale (AUC, 0.99).

A significant advantage of our findings is that the volumetric ratios reported can be automatically extracted within seconds, eliminating the need for time-consuming manual analysis by neuroradiologists. Given the widespread availability of CT scans and their ubiquitous use in clinical routine, the potential for automated screening tools that enhance and expedite radiological assessments is highly promising and is therefore positioned to have an immediate/substantial clinical impact.

Another interesting finding of our study is that the volumetric correlations between automatically and manually derived CT-VCSF volumes in pre-shunt (*ρ* = 0.93, *p*<0.001) and post-shunt procedure (*ρ* = 0.92, *p*<0.001) were similar. This implied that the presence of intraventricular catheters in CT scans had minimal impact on the model’s segmentation accuracy, as shown in **Figure 4**. This consistency underscores the potential for using automated CT ventricular volumes in the ongoing monitoring of iNPH patients after shunt surgery. Such tools could significantly aid in assessing shunt functionality and managing complications related to over- or under-drainage in iNPH and other forms of hydrocephalus. Given that the model performs well even with the presence of shunt catheters in postoperative scans, it could potentially be used to monitor small volumetric changes associated with shunt dysfunction. Additionally, in elderly patients, enlarged ventricles may represent either hydrocephalus or general brain atrophy^51^, which diminishes the specificity of traditional metrics like the Evans’ index. Utilizing the specific ratio of brain volume to intracranial volume (BV/ICV) may yield results that are indicative of generalized atrophy, while the ventricular volume to CSF volume (VCSF/CSF) ratio provides a direct assessment of hydrocephalus, highlighting the potential of such an approach as a next-generation screening tool in clinical settings.

To better integrate this method into clinical settings, further research is needed to explore the relationship between CTVMs and established iNPH metrics such as the Evans’ index and callosal angle. We also plan to explore the potential of these variables for differential diagnosis in conditions like iNPH, dementia, and/or other neurodegenerative diseases. Additionally, we aim to optimize the model’s architecture and hyperparameters and conduct a visual comparison of manual and automated CT-VCSF maps. By involving expert raters (senior neuroradiologists), we hope to identify the sources of systematic bias, further refine our models, and develop generalizable models.

In summary, CTVMs enhanced through deep learning, demonstrate a strong correlation with established clinical indicators of iNPH. Significantly, these measures effectively distinguish iNPH patients from control groups. As such we contend that automated techniques for generating CTVMs and related ratios could serve as valuable tools for identifying hydrocephalus in clinical CT scans. Such methods not only support the diagnosis of iNPH but also offer a practical and cost-effective alternative to MRI and conventional visual rating scales.

## Data Availability

The Gothenburg H70 Birth cohort, Uppsala University Hospital, the Universitätsmedizin Rostock, Germany, and the University of Alabama at Birmingham, United States cannot openly share data according to existing ethical and data sharing approvals, however, relevant data can and will be shared with research groups after submitting a research proposal which has to be approved by the respective study coordinators.

## Acknowledgements

The authors thank Felicia Forseni Flodin, who together with W.S and D.F performed manual segmentations. S.K. is financed by grants from the Swedish state under the agreement between the Swedish government and the county councils, the ALF-agreement (ALFGBG-965923, ALFGBG-81392, ALF GBG-771071), the Alzheimerfonden (AF-842471, AF-737641, AF-929959, AF-939825), the Swedish Research Council (2019-02075), Stiftelsen Psykiatriska Forskningsfonden, Stiftelsen Demensfonden, Stiftelsen Hjalmar Svenssons Forskningsfond, Stiftelsen Wilhelm och Martina Lundgrens vetenskapsfond. J.V. and D.F. are supported by the Swedish Society for Medical Research (SSMF; SG-22-0192-H-01 and PD21-0136). D.F. is also supported by Hjärnfonden (PS2021-0026). Image analysis computations were in part carried out with resources provided by the Swedish National Infrastructure for Computing (SNIC), partially funded by the Swedish Research Council through grant agreement no. 2018-05973. M.Sc. receives funding from the Knut and Alice Wallenberg Foundation (Wallenberg Centre for Molecular and Translational Medicine; KAW2014.0363 and KAW2023.0371), the Swedish Research Council (2017-02869, 2021-02678, 2021-06545 and 2023-06188), the European Union’s Horizon Europe research and innovation program under grant agreement no 101132933 (AD-RIDDLE) and 101112145 (PROMINENT), the National Institute of Health (R01 AG081394-01), Gates Ventures, the National Research Foundation of Korea (RS-2023-00263612), the Swedish state under the agreement between the Swedish government and the County Councils, the ALF-agreement (ALFGBG-813971 and ALFGBG-965326), the Swedish Brain Foundation (FO2021-0311), the Swedish Alzheimer Foundation (AF-994900), the Sahlgrenska Academy at the University of Gothenburg, the Västra Götaland Region R&D (VGFOUREG-995510) and Innovation platforms, Sahlgrenska Science Park and the National Institute for Health and Care Research University College London Hospitals Biomedical Research Centre.

## References

1. Miskin N, Patel H, Franceschi AM, et al. Diagnosis of normal-pressure hydrocephalus: use of traditional measures in the era of volumetric MR imaging. Radiology. 2017;285(1):197–205.

2. Nakajima M, Yamada S, Miyajima M, et al. Guidelines for Management of Idiopathic Normal Pressure Hydrocephalus (Third Edition): Endorsed by the Japanese Society of Normal Pressure Hydrocephalus. Neurologia medico-chirurgica. 2021;61(2):63–97. doi:10.2176/nmc.st.2020-0292

3. Tullberg M, Toma AK, Yamada S, et al. Classification of Chronic Hydrocephalus in Adults: A Systematic Review and Analysis. World Neurosurg. 2024;183:113–122. doi:10.1016/j.wneu.2023.12.094

4. Luciano M, Holubkov R, Williams MA, et al. Placebo-Controlled Effectiveness of Idiopathic Normal Pressure Hydrocephalus Shunting: A Randomized Pilot Trial. Neurosurgery. 2023;92(3):481–489. doi:10.1227/neu.0000000000002225

5. Constantinescu C, Wikkelsø C, Westman E, et al. Prevalence of Possible Idiopathic Normal Pressure Hydrocephalus in Sweden. Neurology. 2024;102(2):e208037. doi:10.1212/WNL.0000000000208037

6. Andersson J, Rosell M, Kockum K, Lilja-Lund O, Söderström L, Laurell K. Prevalence of idiopathic normal pressure hydrocephalus: A prospective, population-based study. PLoS One. 2019;14(5):e0217705. doi:10.1371/journal.pone.0217705

7. Virhammar J, Cesarini KG, Laurell K. The CSF tap test in normal pressure hydrocephalus: evaluation time, reliability and the influence of pain. European Journal of Neurology. 2012;19(2):271–276. doi:10.1111/j.1468-1331.2011.03486.x

8. Williams MA, Malm J. Diagnosis and Treatment of Idiopathic Normal Pressure Hydrocephalus. Continuum (Minneap Minn). 2016;22(2 Dementia):579–599. doi:10.1212/CON.0000000000000305

9. Reis AE, Spano M, Davis-Hayes C, Salama GR. Lumbar Puncture Complications: A Review of Current Literature. Curr Pain Headache Rep. Published online May 22, 2024. doi:10.1007/s11916-024-01262-2

10. Zhou X, Xia J. Application of Evans Index in Normal Pressure Hydrocephalus Patients: A Mini Review. Frontiers in Aging Neuroscience. 2022;13. Accessed June 3, 2022. https://www.frontiersin.org/article/10.3389/fnagi.2021.783092

11. Ishii K, Kanda T, Harada A, et al. Clinical impact of the callosal angle in the diagnosis of idiopathic normal pressure hydrocephalus. Eur Radiol. 2008;18(11):2678–2683. doi:10.1007/s00330-008-1044-4

12. Virhammar J, Laurell K, Cesarini KG, Larsson EM. The callosal angle measured on MRI as a predictor of outcome in idiopathic normal-pressure hydrocephalus. J Neurosurg. 2014;120(1):178–184. doi:10.3171/2013.8.JNS13575

13. Craven CL, Toma AK, Mostafa T, Patel N, Watkins LD. The predictive value of DESH for shunt responsiveness in idiopathic normal pressure hydrocephalus. J Clin Neurosci. 2016;34:294–298. doi:10.1016/j.jocn.2016.09.004

14. Kockum K, Lilja-Lund O, Larsson EM, et al. The idiopathic normal-pressure hydrocephalus Radscale: a radiological scale for structured evaluation. European Journal of Neurology. 2018;25(3):569–576. doi:10.1111/ene.13555

15. Fällmar D, Andersson O, Kilander L, Löwenmark M, Nyholm D, Virhammar J. Imaging features associated with idiopathic normal pressure hydrocephalus have high specificity even when comparing with vascular dementia and atypical parkinsonism. Fluids and Barriers of the CNS. 2021;18(1):35. doi:10.1186/s12987-021-00270-3

16. Moore DW, Kovanlikaya I, Heier LA, et al. A Pilot Study of Quantitative MRI Measurements of Ventricular Volume and Cortical Atrophy for the Differential Diagnosis of Normal Pressure Hydrocephalus. Neurol Res Int. 2012;2012:718150. doi:10.1155/2012/718150

17. Neikter J, Agerskov S, Hellström P, et al. Ventricular Volume Is More Strongly Associated with Clinical Improvement Than the Evans Index after Shunting in Idiopathic Normal Pressure Hydrocephalus. AJNR Am J Neuroradiol. 2020;41(7):1187–1192. doi:10.3174/ajnr.A6620

18. Cogswell PM, Murphy MC, Senjem ML, et al. Changes in Ventricular and Cortical Volumes following Shunt Placement in Patients with Idiopathic Normal Pressure Hydrocephalus. American Journal of Neuroradiology. Published online October 21, 2021. doi:10.3174/ajnr.A7323

19. Virhammar J, Laurell K, Cesarini KG, Larsson EM. Increase in callosal angle and decrease in ventricular volume after shunt surgery in patients with idiopathic normal pressure hydrocephalus. Journal of Neurosurgery. 2018;130(1):130–135. doi:10.3171/2017.8.JNS17547

20. Kockum K, Virhammar J, Riklund K, Söderström L, Larsson EM, Laurell K. Standardized image evaluation in patients with idiopathic normal pressure hydrocephalus: consistency and reproducibility. Neuroradiology. 2019;61(12):1397–1406. doi:10.1007/s00234-019-02273-2

21. Ntiri EE, Holmes MF, Forooshani PM, et al. Improved Segmentation of the Intracranial and Ventricular Volumes in Populations with Cerebrovascular Lesions and Atrophy Using 3D CNNs. Neuroinform. 2021;19(4):597–618. doi:10.1007/s12021-021-09510-1

22. Quon JL, Han M, Kim LH, et al. Artificial intelligence for automatic cerebral ventricle segmentation and volume calculation: a clinical tool for the evaluation of pediatric hydrocephalus. Journal of Neurosurgery: Pediatrics. 2020;27(2):131–138. doi:10.3171/2020.6.PEDS20251

23. Dubost F, Bruijne M de, Nardin M, et al. Multi-atlas image registration of clinical data with automated quality assessment using ventricle segmentation. Medical Image Analysis. 2020;63:101698. doi:10.1016/j.media.2020.101698

24. Shao M, Han S, Carass A, et al. Brain ventricle parcellation using a deep neural network: Application to patients with ventriculomegaly. NeuroImage: Clinical. 2019;23:101871. doi:10.1016/j.nicl.2019.101871

25. Oike R, Inoue Y, Matsuzawa K, Sorimachi T. Screening for idiopathic normal pressure hydrocephalus in the elderly after falls. Clinical Neurology and Neurosurgery. 2021;205:106635. doi:10.1016/j.clineuro.2021.106635

26. Jyothi P, Singh AR. Deep learning models and traditional automated techniques for brain tumor segmentation in MRI: a review. Artif Intell Rev. 2023;56(4):2923–2969. doi:10.1007/s10462-022-10245-x

27. Akkus Z, Galimzianova A, Hoogi A, Rubin DL, Erickson BJ. Deep learning for brain MRI segmentation: state of the art and future directions. Journal of digital imaging. 2017;30(4):449–459.

28. Kadaba Sridhar S, Kuang R, Dysterheft Robb J, Samadani U. A ventriculomegaly feature computational pipeline to improve the screening of normal pressure hydrocephalus on CT. J Neurosurg. Published online March 8, 2024:1–11. doi:10.3171/2023.12.JNS231780

29. Kaipainen AL, Pitkänen J, Haapalinna F, et al. A novel CT-based automated analysis method provides comparable results with MRI in measuring brain atrophy and white matter lesions. Neuroradiology. 2021;63(12):2035–2046. doi:10.1007/s00234-021-02761-4

30. Huff TJ, Ludwig PE, Salazar D, Cramer JA. Fully automated intracranial ventricle segmentation on CT with 2D regional convolutional neural network to estimate ventricular volume. Int J CARS. 2019;14(11):1923–1932. doi:10.1007/s11548-019-02038-5

31. Cai JC, Akkus Z, Philbrick KA, et al. Fully Automated Segmentation of Head CT Neuroanatomy Using Deep Learning. Radiology: Artificial Intelligence. 2020;2(5):e190183. doi:10.1148/ryai.2020190183

32. Zhou X, Ye Q, Yang X, et al. AI-based medical e-diagnosis for fast and automatic ventricular volume measurement in patients with normal pressure hydrocephalus. Neural Comput Appl. Published online February 24, 2022:1–10. doi:10.1007/s00521-022-07048-0

33. Cherukuri V, Ssenyonga P, Warf BC, Kulkarni AV, Monga V, Schiff SJ. Learning Based Segmentation of CT Brain Images: Application to Postoperative Hydrocephalic Scans. IEEE Transactions on Biomedical Engineering. 2018;65(8):1871–1884. doi:10.1109/TBME.2017.2783305

34. Srikrishna M, Pereira JB, Heckemann RA, et al. Deep learning from MRI-derived labels enables automatic brain tissue classification on human brain CT. NeuroImage. 2021;244:118606. doi:10.1016/j.neuroimage.2021.118606

35. Azizpour H, Razavian AS, Sullivan J, Maki A, Carlsson S. Factors of Transferability for a Generic ConvNet Representation. IEEE Trans Pattern Anal Mach Intell. 2016;38(9):1790–1802. doi:10.1109/TPAMI.2015.2500224

36. Kim HE, Cosa-Linan A, Santhanam N, Jannesari M, Maros ME, Ganslandt T. Transfer learning for medical image classification: a literature review. BMC Medical Imaging. 2022;22(1):69. doi:10.1186/s12880-022-00793-7

37. Rydberg Sterner T, Ahlner F, Blennow K, et al. The Gothenburg H70 Birth cohort study 2014–16: design, methods and study population. European journal of epidemiology. 2019;34(2):191–209. doi:10.1007/s10654-018-0459-8

38. Fischl B. FreeSurfer. Neuroimage. 2012;62(2):774–781.

39. Muehlboeck JS, Westman E, Simmons A. TheHiveDB image data management and analysis framework. Frontiers in Neuroinformatics. 2013;7. doi:10.3389/fninf.2013.00049

40. Ashburner J, Friston KJ. Rigid body registration. Statistical parametric mapping: The analysis of functional brain images. Published online 2007:49–62.

41. Perez L, Wang J. The Effectiveness of Data Augmentation in Image Classification Using Deep Learning. arXiv; 2017. doi:10.48550/arXiv.1712.04621

42. Dartora C, Marseglia A, Mårtensson G, et al. A deep learning model for brain age prediction using minimally preprocessed T1w images as input. Front Aging Neurosci. 2024;15:1303036. doi:10.3389/fnagi.2023.1303036

43. Pyrgelis ES, Velonakis G, Papageorgiou SG, Stefanis L, Kapaki E, Constantinides VC. Imaging Markers for Normal Pressure Hydrocephalus: An Overview. Biomedicines. 2023;11(5):1265. doi:10.3390/biomedicines11051265

44. Thavarajasingam SG, El-Khatib M, Vemulapalli K, et al. Radiological predictors of shunt response in the diagnosis and treatment of idiopathic normal pressure hydrocephalus: a systematic review and meta-analysis. Acta Neurochir (Wien). 2023;165(2):369–419. doi:10.1007/s00701-022-05402-8

45. He W, Fang X, Wang X, et al. A new index for assessing cerebral ventricular volume in idiopathic normal-pressure hydrocephalus: a comparison with Evans’ index. Neuroradiology. 2020;62(6):661–667. doi:10.1007/s00234-020-02361-8

46. Yamada S, Ishikawa M, Yamamoto K. Optimal Diagnostic Indices for Idiopathic Normal Pressure Hydrocephalus Based on the 3D Quantitative Volumetric Analysis for the Cerebral Ventricle and Subarachnoid Space. American Journal of Neuroradiology. 2015;36(12):2262–2269. doi:10.3174/ajnr.A4440

47. Ryska P, Slezak O, Eklund A, Malm J, Salzer J, Zizka J. Radiological markers of idiopathic normal pressure hydrocephalus: Relative comparison of their diagnostic performance. Journal of the Neurological Sciences. 2020;408:116581. doi:10.1016/j.jns.2019.116581

48. Kockum K, Lilja-Lund O, Larsson EM, et al. The idiopathic normal-pressure hydrocephalus Radscale: a radiological scale for structured evaluation. European Journal of Neurology. 2018;25(3):569–576. doi:10.1111/ene.13555

49. Srikrishna M, Ashton NJ, Moscoso A, et al. CT-based volumetric measures obtained through deep learning: Association with biomarkers of neurodegeneration. Alzheimer’s & Dementia. 2024;20(1):629–640.

